# Laboratory findings, signs and symptoms, clinical outcomes of Patients with COVID-19 Infection: an updated systematic review and meta-analysis

**DOI:** 10.1101/2020.03.25.20043703

**Authors:** Mina Ebrahimi, Amal Saki Malehi, Fakher Rahim

## Abstract

**Background and Aim:** Coronaviruses disease 2019 (COVID-19), for the first time detected in Wuhan, China, rapidly speared around the world and be a Public Health Emergency of International Concern (PHEIC). The aim of the current survey is collecting laboratory findings, analysis them and reporting a specific pattern for help to COVID-19 diagnosis.

**Methods:** To collect laboratory characteristics, we searched “PubMed” electronic database with the following keywords: “COVID-19” “2019 novel coronavirus” “laboratory findings” “clinical characteristics”.

**Results:** Once the initial searches 493 studies were yielded. After removing duplicates studies 480 studies were remained. The 12 studies obtained from the literature, of which 58.3% (7) of studies were case-control (8–14), and 41.7% (5) remaining studies were designed as cross-sectional (1,15–18)

**Conclusion:** The result of the current study showed that in the early stage of COVID-19 infection, maybe there are not significant laboratory findings, but with disease progression, the one or more than signs include increasing AST, ALT, LDH, CK, CRP, ESR, WBC, neutrophil, and decreasing Hemoglobin, lymphocyte count, eosinophil count can be seen. Elevating D-dimer and FDP are associated with ARDS development and can be used as prognostic factors.

## Introduction

Coronaviruses disease 2019 (COVID-19), for the first time detected in Wuhan, China, rapidly speared around the world and be a Public Health Emergency of International Concern (PHEIC). COVID-19 by involving the respiratory system can rapidly develop to the acute respiratory distress syndrome (1). Genetic studies revealed that COVID-19 has 1 88% similarity with severe acute respiratory syndrome (SARS) and Middle East respiratory syndrome (MERS) (2). Despite to SARS, COVID-19 has longer incubation time, lack of pathologic symptoms, and transferability in the incubation period made it more serious pathogenic and rapidly transmission (3). Reverse real-time PCR (rRT-PCR) used for specific diagnosis of COVID-19, but like the other laboratories tests, has a false-negative result; specially in the early stage of disease.

Nevertheless, a specific laboratory diagnosis along with other clinical characteristics and their association with severity of the disease is necessary. Several studies reported different laboratory findings (4–6). The aim of the current survey is collecting laboratory findings, analysis them and reporting a specific pattern for help to COVID-19 diagnosis.

## Methods

### Search strategy

To collect laboratory characteristics, we searched “PubMed” electronic database with the following keywords: “COVID-19” “2019 novel coronavirus” “laboratory findings” “clinical characteristics”. There was no restriction for date and language. The present survey was performed base on PRISMA-P guideline Preferred reporting items for the systematic review and meta-analysis protocols (PRISMA-P) 2015 statement (7).

### Study selection

Two reviewers (M.E and F.R) screened all studies first by title and abstract, then reviewed the full text of the article. The inclusion criteria for selecting studies were as follows; evaluating hematological, coagulation, biochemistry, and serological laboratory tests. Exclusion criteria were considered as studies with just molecular reports and studies that report laboratory results with the percentage.

### Data extraction

Two independent reviewers (F.R and M.E) extract the data from included studies. Data were extracted by the following key characteristics: author, publication year, country, type of study, sample size, result and final finding.

### Statistical Analysis of Data

Cochran Chi-square test and I^2^ were used to assessing heterogeneity among studies. A fixed-effects model was used When I^2^ < 50%, while when I^2^> 50%, a random-effects model was selected. If there was statistical heterogeneity among the results, a further sensitivity analysis was conducted to determine the source of heterogeneity. After the significant clinical heterogeneity was excluded, the randomized effects model was used for meta-analysis. Publication bias was evaluated using the Funnel plot and Egger test. P < 0.05 was considered as statistical significance (2-sided). All data were analyzed using the STAT 15 software.

## Result

Once the initial searches 493 studies were yielded. After removing duplicates studies 480 studies were remained. One reviewer (M.E) screened 480 articles by title and abstract. 449 studies were excluded based on exclusion criteria. After evaluating the full text of studies by inclusion and exclusion criteria, 12 articles remained (Figure 1).

**Figure 1.**
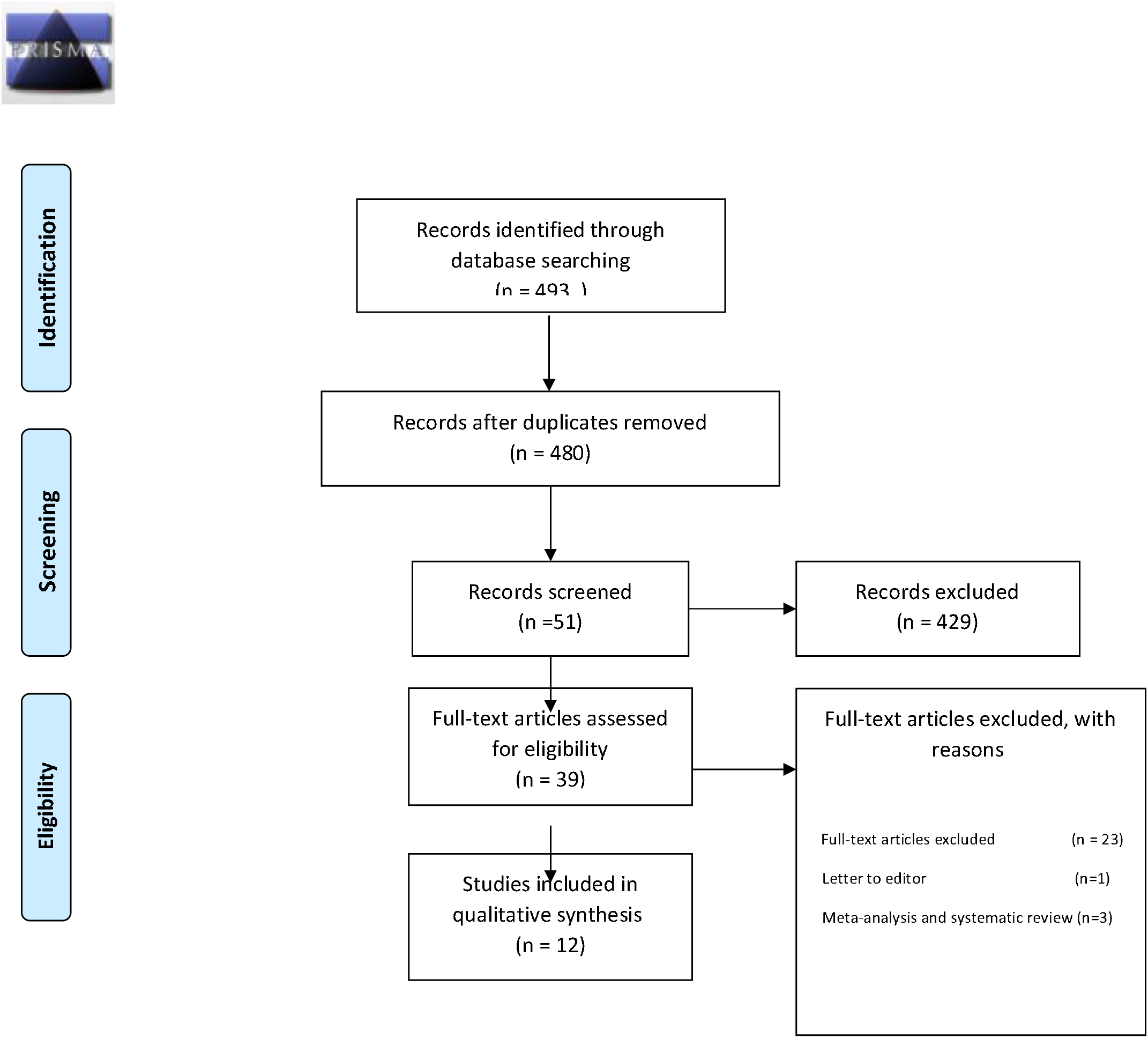
PRISMA flow diagram of the literature search

### Study characteristics and quality assessment

The 12 studies obtained from the literature systematic review are presented in Table 1. Of these, 58.3% (7) of studies were case-control (8–14), and 41.7% (5) remaining studies were designed as cross-sectional (1,15–18). All researches conducted in China. There is one study that investigated the association between coagulation abnormalities with prognosis in infected patients with COVID-19 (8).

**Table 1.**
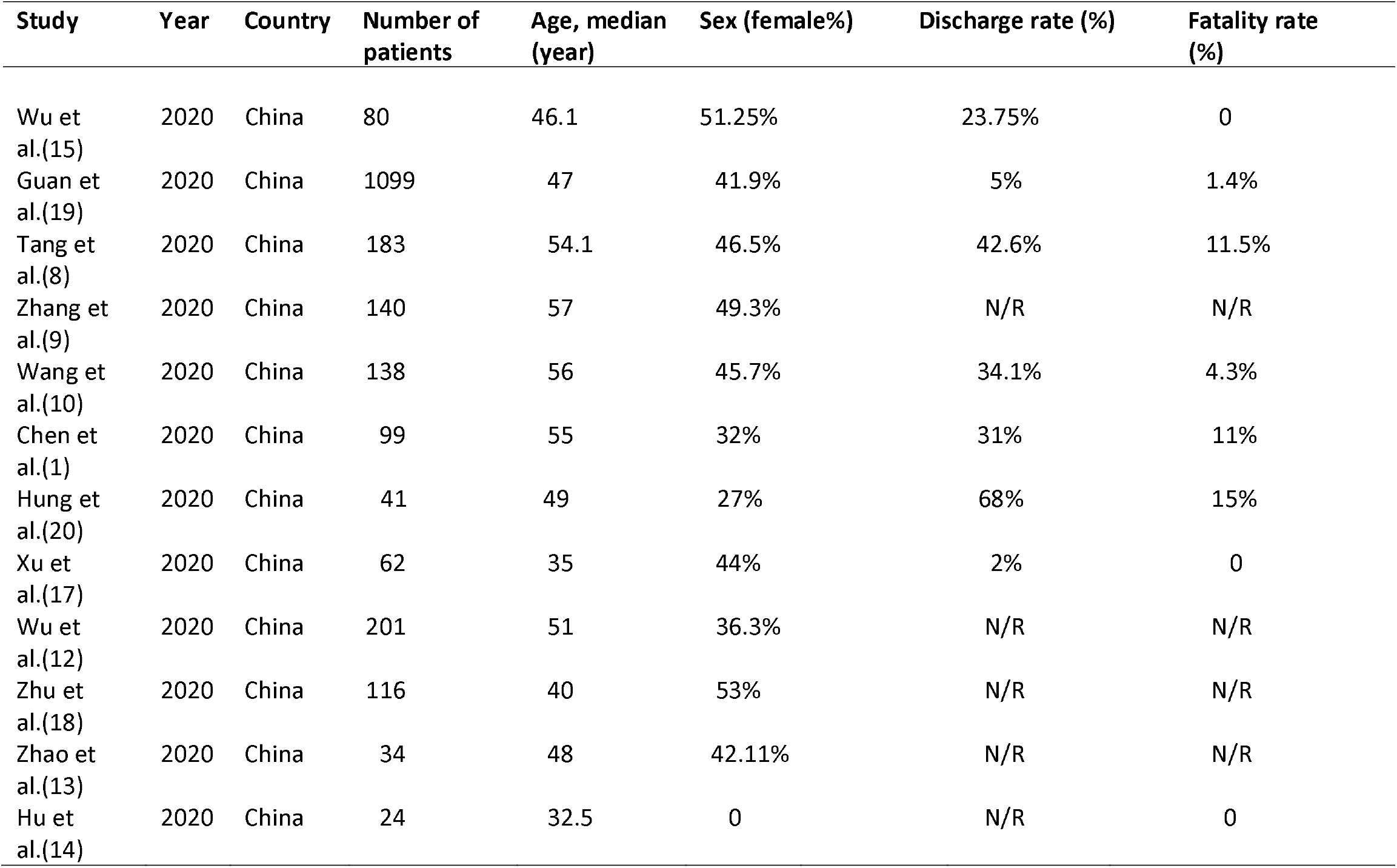
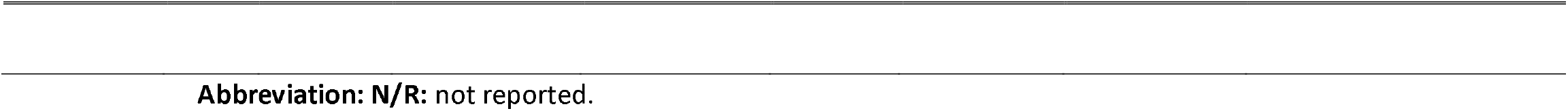
Demographics of the Included Studies

| Study | Year | Country | Number of patients | Age, median (year) | Sex (female%) | Discharge rate (%) | Fatality rate (%) |
| --- | --- | --- | --- | --- | --- | --- | --- |
| Wu et al.(15) | 2020 | China | 80 | 46.1 | 51.25% | 23.75% | 0 |
| Guan et al.(19) | 2020 | China | 1099 | 47 | 41.9% | 5% | 1.4% |
| Tang et al.(8) | 2020 | China | 183 | 54.1 | 46.5% | 42.6% | 11.5% |
| Zhang et al.(9) | 2020 | China | 140 | 57 | 49.3% | N/R | N/R |
| Wang et al.(10) | 2020 | China | 138 | 56 | 45.7% | 34.1% | 4.3% |
| Chen et al.(1) | 2020 | China | 99 | 55 | 32% | 31% | 11% |
| Hung et al.(20) | 2020 | China | 41 | 49 | 27% | 68% | 15% |
| Xu et al.(17) | 2020 | China | 62 | 35 | 44% | 2% | 0 |
| Wu et al.(12) | 2020 | China | 201 | 51 | 36.3% | N/R | N/R |
| Zhu et al.(18) | 2020 | China | 116 | 40 | 53% | N/R | N/R |
| Zhao et al.(13) | 2020 | China | 34 | 48 | 42.11% | N/R | N/R |
| Hu et al.(14) | 2020 | China | 24 | 32.5 | 0 | N/R | 0 |

**Abbreviation: N/R:** not reported.

Just one study had a sample size greater than 1000 (16) (Table 1). The demographic and quality assessment of included studies were shown in Table 1 and Table 2.

**Table 2.** The quality assessment of included studies

| Study | 1 | 2 | 3 | 4 | 5 | 6 | 7 | 8 | Score |
| --- | --- | --- | --- | --- | --- | --- | --- | --- | --- |
| Wu et al.(15) | 2 | 2 | 2 | 2 | 2 | 1 | 2 | 0 | 13 |
| Guan et al.(19) | 2 | 2 | 2 | 2 | 2 | 0 | 0 | 0 | 10 |
| Tang et al.(8) | 2 | 2 | 2 | 2 | 2 | 1 | 2 | 0 | 13 |
| Zhang et al.(9) | 2 | 2 | 2 | 2 | 2 | 0 | 0 | 0 | 10 |
| Wang et al.(10) | 2 | 2 | 2 | 2 | 2 | 1 | 2 | 0 | 13 |
| Chen et al.(1) | 2 | 2 | 2 | 2 | 2 | 1 | 1 | 0 | 12 |
| Hung et al.(20) | 2 | 2 | 2 | 2 | 2 | 1 | 2 | 0 | 13 |
| Xu et al.(17) | 2 | 2 | 2 | 2 | 2 | 1 | 2 | 0 | 13 |
| Wu et al.(12) | 2 | 2 | 2 | 2 | 2 | 1 | 1 | 0 | 12 |
| Zhu et al.(18) | 2 | 2 | 2 | 2 | 2 | 0 | 0 | 0 | 10 |
| Zhao et al.(13) | 2 | 2 | 2 | 2 | 2 | 1 | 2 | 0 | 13 |
| Hu et al.(14) | 2 | 2 | 2 | 2 | 2 | 1 | 2 | 0 | 13 |

①A clearly stated aim; ②Inclusion of consecutive patients; ③Prospective collection of data; ④ Endpoints appropriate to the aim of the study; ⑤Unbiased assessment of the study endpoint; ⑥Follow-up period appropriate to the aim of the study; ⑦Loss to follow up less than 5%; ⑧ Prospective calculation of the study size. The items are scored 0 (not reported), 1 (reported but inadequate) or 2 (reported and adequate). The global ideal score being 16 for non-comparative studies.

The result of laboratory finding analysis showed that, lymphocytopenia, increasing C-reactive protein (CRP) and erythrocyte sedimentation rate (ESR) (Table 3). Egger’s test was done for all laboratory tests and the result showed that there was no publication bias (Table 3). Among these, measuring glucose has the lowest important. The most reported clinical findings were fever, cough, diarrhea, and fatigue; whereas the chest pain and muscle ache were reported just by two studies (Table 4). Egger test results, revealed there was no publication bias (Table 4). After investigating for comorbidity situation analysis, it revealed that patients with chronic obstructive pulmonary disease (COPD), hypertension, cardiovascular disease (CVD), diabetes, Cerebrovascular disease (CBD), malignancy and kidney abnormalities (Table 5). The details of reported results were shown in supplemental 1.

**Table 3.**
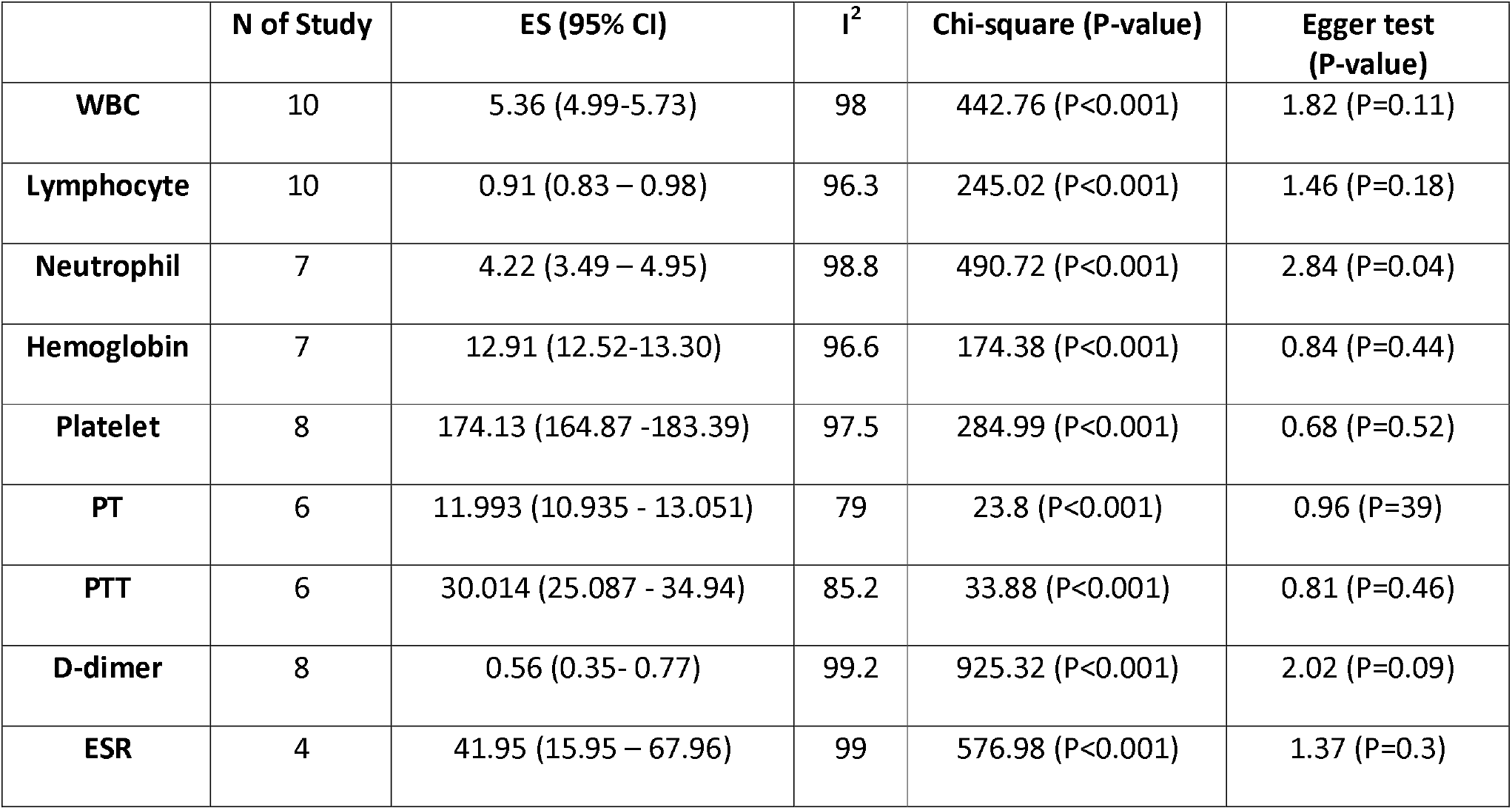
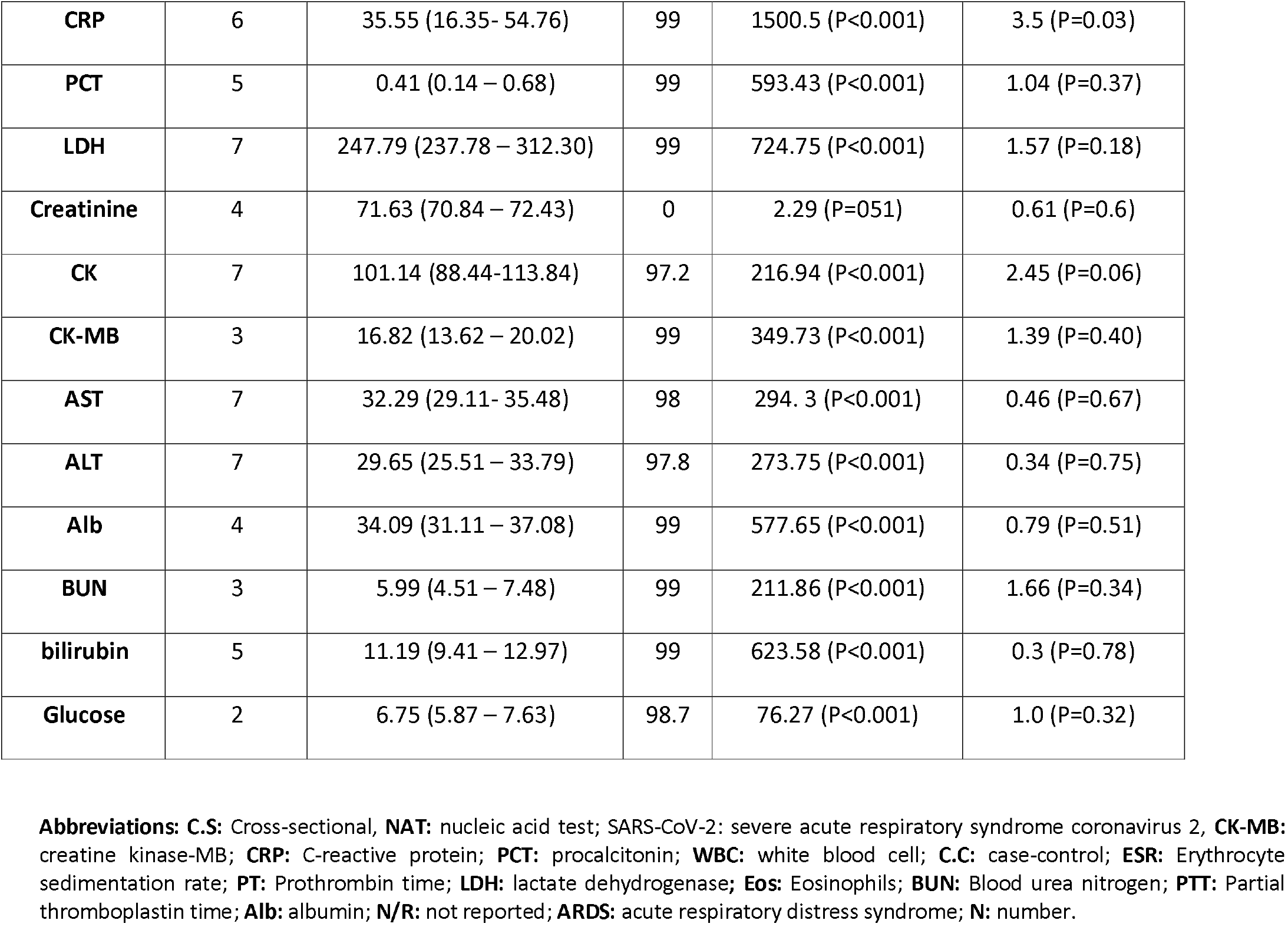
The result of laboratory findings analysis in COVID-19 infected patients.

|  | **N of Study** | **ES (95% CI)** | **I<sup>2</sup>** | **Chi-square (P-value)** | **Egger test (P-value)** |
| --- | --- | --- | --- | --- | --- |
| **WBC** | 10 | 5.36 (4.99-5.73) | 98 | 442.76 (P<0.001) | 1.82 (P=0.11) |
| **Lymphocyte** | 10 | 0.91 (0.83 – 0.98) | 96.3 | 245.02 (P<0.001) | 1.46 (P=0.18) |
| **Neutrophil** | 7 | 4.22 (3.49 – 4.95) | 98.8 | 490.72 (P<0.001) | 2.84 (P=0.04) |
| **Hemoglobin** | 7 | 12.91 (12.52-13.30) | 96.6 | 174.38 (P<0.001) | 0.84 (P=0.44) |
| **Platelet** | 8 | 174.13 (164.87 -183.39) | 97.5 | 284.99 (P<0.001) | 0.68 (P=0.52) |
| **PT** | 6 | 11.993 (10.935 - 13.051) | 79 | 23.8 (P<0.001) | 0.96 (P=0.39) |
| **PTT** | 6 | 30.014 (25.087 - 34.94) | 85.2 | 33.88 (P<0.001) | 0.81 (P=0.46) |
| **D-dimer** | 8 | 0.56 (0.35- 0.77) | 99.2 | 925.32 (P<0.001) | 2.02 (P=0.09) |
| **ESR** | 4 | 41.95 (15.95 – 67.96) | 99 | 576.98 (P<0.001) | 1.37 (P=0.3) |
| **CRP** | 6 | 35.55 (16.35 - 54.76) | 99 | 1500.5 (P<0.001) | 3.5 (P=0.03) |
| **PCT** | 5 | 0.41 (0.14 – 0.68) | 99 | 593.43 (P<0.001) | 1.04 (P=0.37) |
| **LDH** | 7 | 247.79 (237.78 – 312.30) | 99 | 724.75 (P<0.001) | 1.57 (P=0.18) |
| **Creatinine** | 4 | 71.63 (70.84 – 72.43) | 0 | 2.29 (P=0.51) | 0.61 (P=0.6) |
| **CK** | 7 | 101.14 (88.44-113.84) | 97.2 | 216.94 (P<0.001) | 2.45 (P=0.06) |
| **CK-MB** | 3 | 16.82 (13.62 – 20.02) | 99 | 349.73 (P<0.001) | 1.39 (P=0.40) |
| **AST** | 7 | 32.29 (29.11- 35.48) | 98 | 294. 3 (P<0.001) | 0.46 (P=0.67) |
| **ALT** | 7 | 29.65 (25.51 – 33.79) | 97.8 | 273.75 (P<0.001) | 0.34 (P=0.75) |
| **Alb** | 4 | 34.09 (31.11 – 37.08) | 99 | 577.65 (P<0.001) | 0.79 (P=0.51) |
| **BUN** | 3 | 5.99 (4.51 – 7.48) | 99 | 211.86 (P<0.001) | 1.66 (P=0.34) |
| **bilirubin** | 5 | 11.19 (9.41 – 12.97) | 99 | 623.58 (P<0.001) | 0.3 (P=0.78) |
| **Glucose** | 2 | 6.75 (5.87 – 7.63) | 98.7 | 76.27 (P<0.001) | 1.0 (P=0.32) |

**Abbreviations:** **C.S:** Cross-sectional, **NAT:** nucleic acid test; SARS-CoV-2: severe acute respiratory syndrome coronavirus 2, **CK-MB:** creatine kinase-MB; **CRP:** C-reactive protein; **PCT:** procalcitonin; **WBC:** white blood cell; **C.C:** case-control; **ESR:** Erythrocyte sedimentation rate; **PT:** Prothrombin time; **LDH:** lactate dehydrogenase; **Eos:** Eosinophils; **BUN:** Blood urea nitrogen; **PTT:** Partial thromboplastin time; **Alb:** albumin; **N/R:** not reported; **ARDS:** acute respiratory distress syndrome; **N:** number.

**Table 4.** The result of Symptoms analysis in COVID-19 infected patients.

|  | **N of Study** | **ES (95% CI)** | **I<sup>2</sup>** | **Chi-square (P-value)** | **Egger test (P-value)** |
| --- | --- | --- | --- | --- | --- |
| **Fever** | 11 | 0.74 (0.56- 0.89) | 98 | 576.69 (P<0.001)) | 2.05 (P=0.07) |
| **Cough** | 11 | 0.63 (0.54 – 0.71) | 91 | 111.64 (P<0.001) | 1.06 (P=0.32) |
| **Fatigue** | 9 | 0.36 (0.24 – 0.49) | 95.4 | 172.31 (P<0.001) | 0.12 (P=0.91) |
| **Muscle ache** | 2 | 0.16 (0.11 – 0.22) | 0 | ----- | 1.0 (P=0.32) |
| **Headache and mental disorder** | 8 | 0.11 (0.06 – 0.16) | 84.4 | 44.92 (P<0.001) | 0.37 (P=0.73) |
| **Sore throat** | 3 | 0.11 (0.06 – 0.17) | 74.7 | 7.89 (P=0.02) | 0.91 (P=0.53) |
| **Chest pain** | 2 | 0.03 (0.01- 0.06) | 0 | ---- | 1.0 (P=0.32) |
| **Diarrhea** | 9 | 0.04 (0.02 – 0.07) | 69.8 | 26.52 (P<0.001) | 0.34 (P=0.74) |
| **Nausea and vomiting** | 5 | 0.06 (0.02 – 0.11) | 88.5 | 34.62 (P<0.001) | 0.47 (P=0.67) |

**Abbreviation: N:** number

**Table 5.** The result of comorbidity analysis in COVID-19 infected patients.

|  | **N of Study** | **ES (95% CI)** | **I<sup>2</sup>** | **Chi-square (P-value)** | **Egger test (P-value)** |
| --- | --- | --- | --- | --- | --- |
| **Hypertension** | 9 | 0.15 (0.10 – 0.21) | 85.4 | 54.83 (P<0.001)) | 0.01 (P=0.99) |
| **CVD** | 8 | 0.08 (0.03 – 0.14) | 92 | 86.76 (P<0.001) | 1.7 (P=0.14) |
| **Diabetes** | 8 | 0.08 (0.05 – 0.11) | 65.9 | 20.51 (P<0.001) | 0.69 (P=0.51) |
| **Malignancy** | 8 | 0.01 (0.0 – 0.03) | 61.5 | 18.02 (P=0.01) | 0.75 (P=0.48) |
| **CBD** | 8 | 0.02 (0.01 – 0.03) | 35.32 | 10.82 (P=0.15) | 0.95 (P=.038) |
| **COPD** | 10 | 0.01 (0.0 – 0.02) | 0 | 7.18 (P=0.62) | 0.66 (P=0.53) |
| **kidney** | 8 | 0.01 (0.0 – 0.02) | 25.82 | 9.44 (P=0.22) | 1.38 (P=0.17) |
| **Liver** | 7 | 0.04 (0.03 – 0.06) | 31.61 | 8.77 (P=0.19) | 0.48 (P=0.65) |
| **Gastrointestinal** | 3 | 0.06 (0.04 – 0.09) | 52.34 | 4.20 (P=0.12) | 0.09 (P=0.94) |
| **Endocrine** | 4 | 0.05 (0.01 – 0.11) | 84.92 | 19.89 (P<0.001) | 1.69 (P=0.23) |
| **HIV** | 2 | 0.01 (0.0 – 0.02) | 0 | ---- | 1.0 (P=0.32) |

**Abbreviations:** CVD: Cardiovascular disease; CBD: Cerebrovascular disease; COPD: chronic obstructive pulmonary disease, N: number.

### Clinical Findings

**Table 6.** Result of clinical outcome analysis

|  | **N of Study** | **ES (95% CI)** | **I<sup>2</sup>** | **Chi-square (P-value)** | **Egger test (P-value)** |
| --- | --- | --- | --- | --- | --- |
| **Remained in hospital** | 8 | 0.56 (0.27- 0.83) | 99.2 | 1014.66 (P<0.001)) | 1.85 (P=0.11) |
| **Discharged** | 9 | 0.33 (0.13 – 0.56) | 98.7 | 618 (P<0.001) | 1.88 (P=0.10) |
| **Died** | 9 | 0.05 (0.01 – 0.11) | 94.29 | 140.16 (P<0.001) | 1.01 (P=0.35) |

**Abbreviation: N: number**

**Figure 1.**
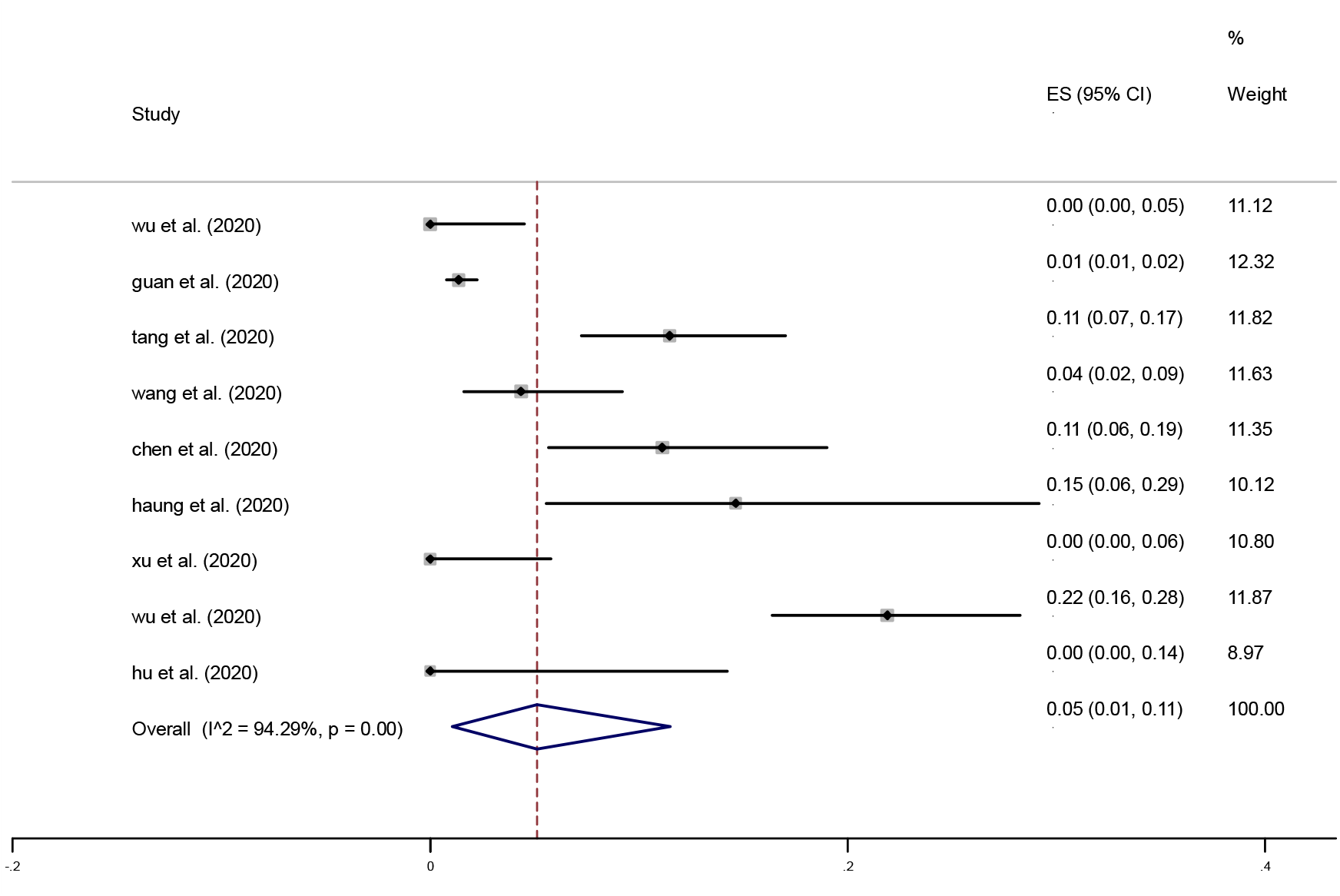
Forest Plot of died proportion

**Figure 1.**
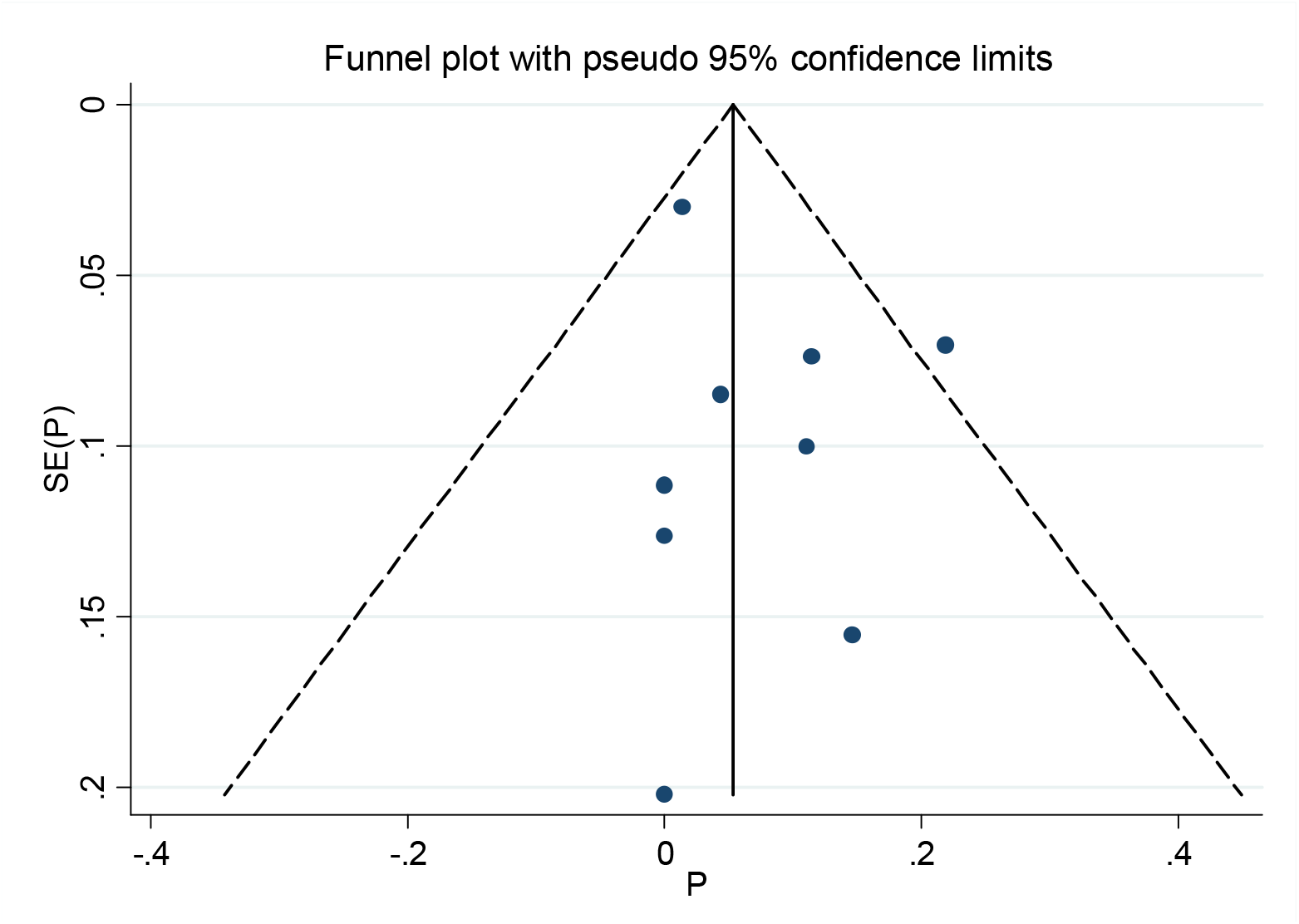
Funnel plot for died proportion

**Figure 2.**
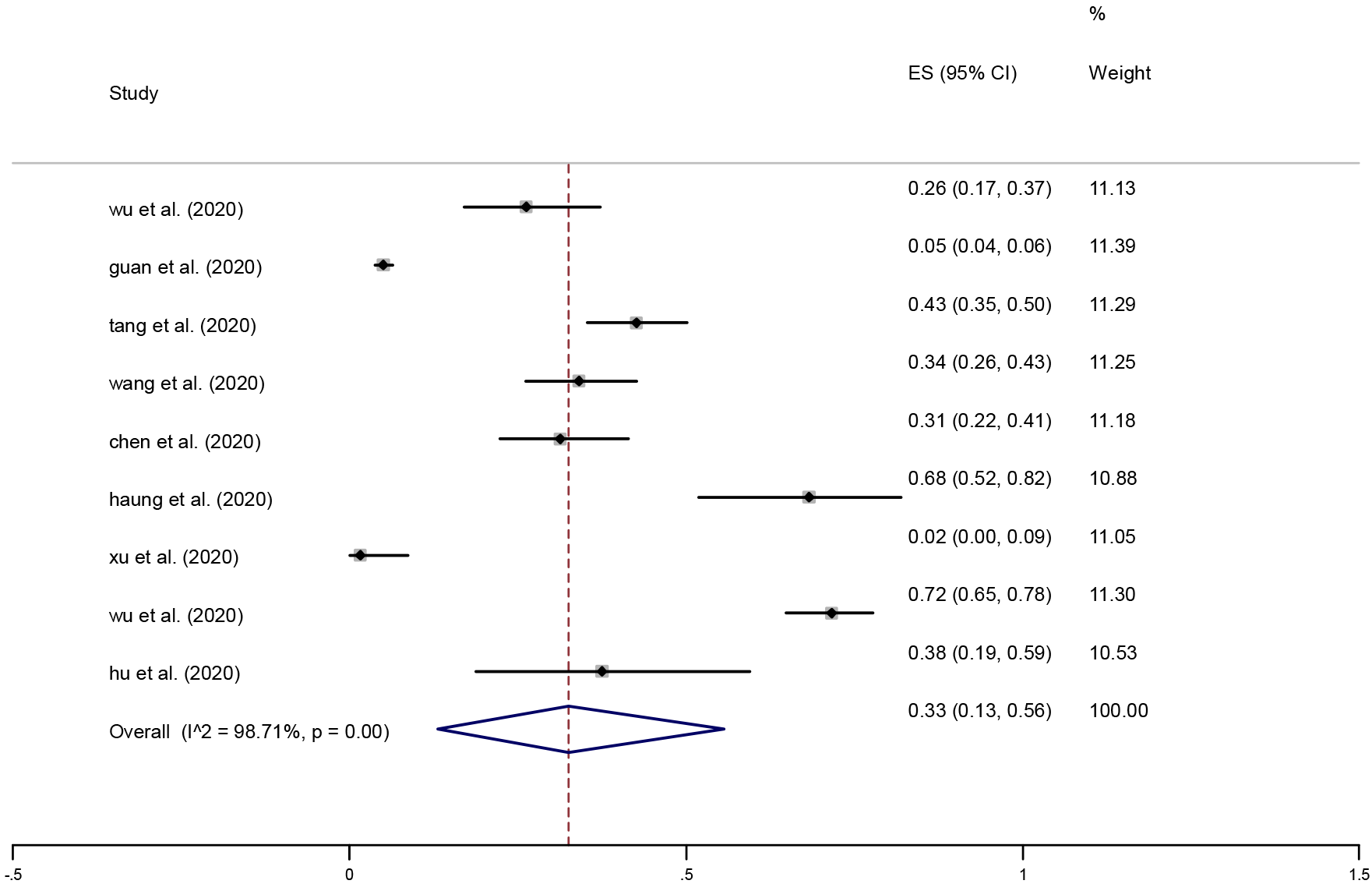
Forest plot of discharged proportion

**Figure 3.**
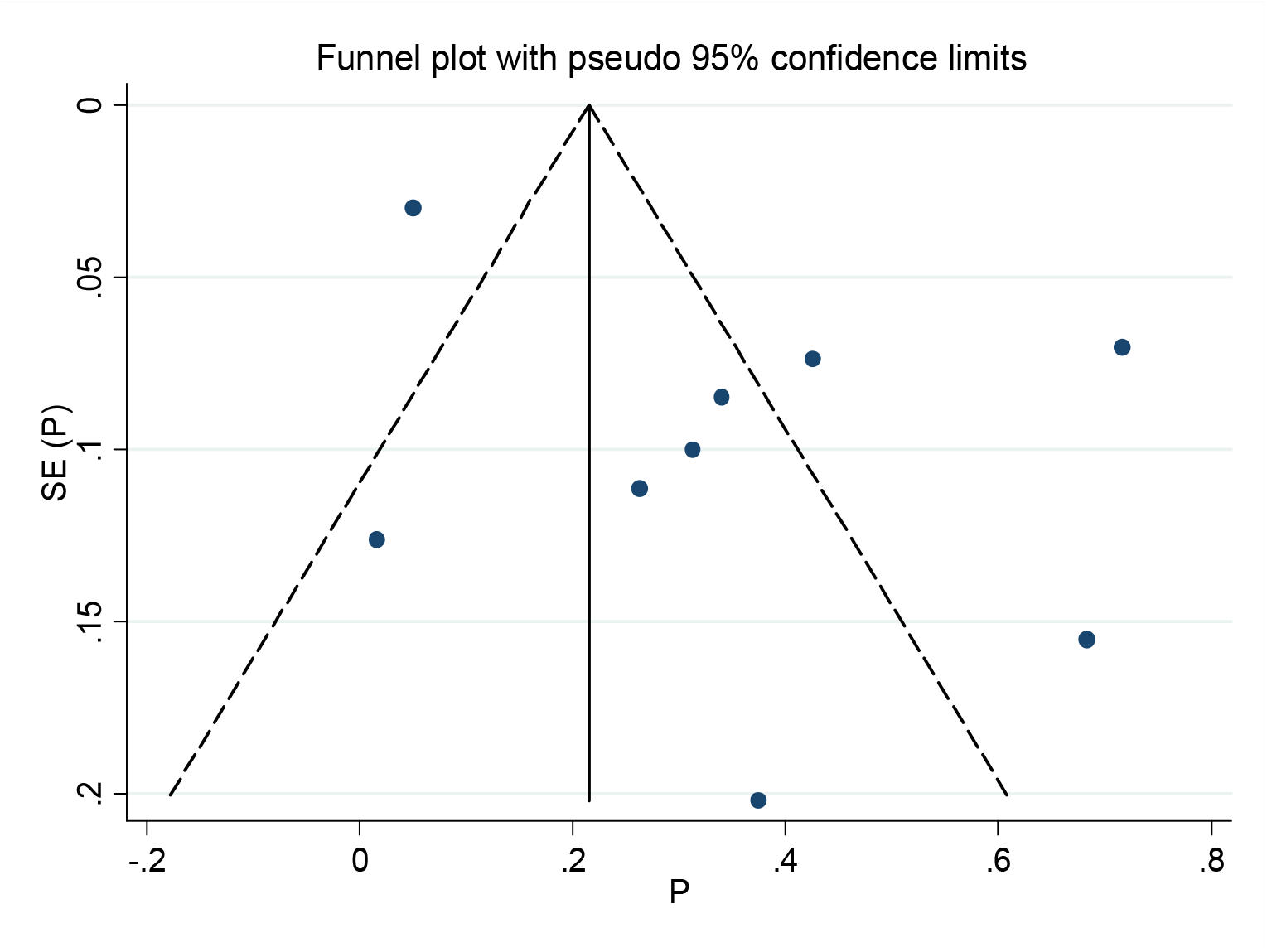
Funnel plot of discharged proportion

**Figure 4.**
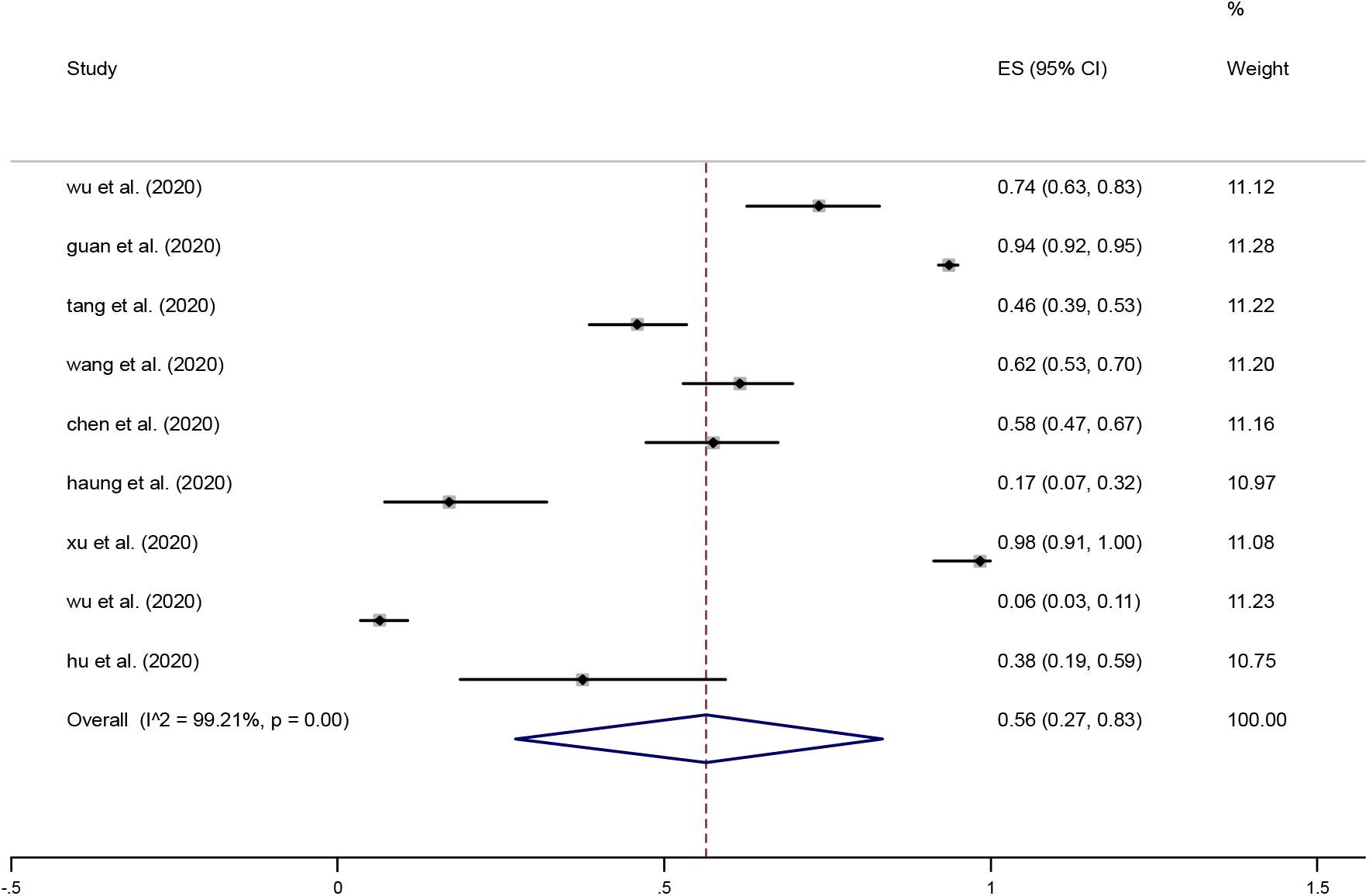
Forest plot for remain in hospital proportion

**Figure 5.**
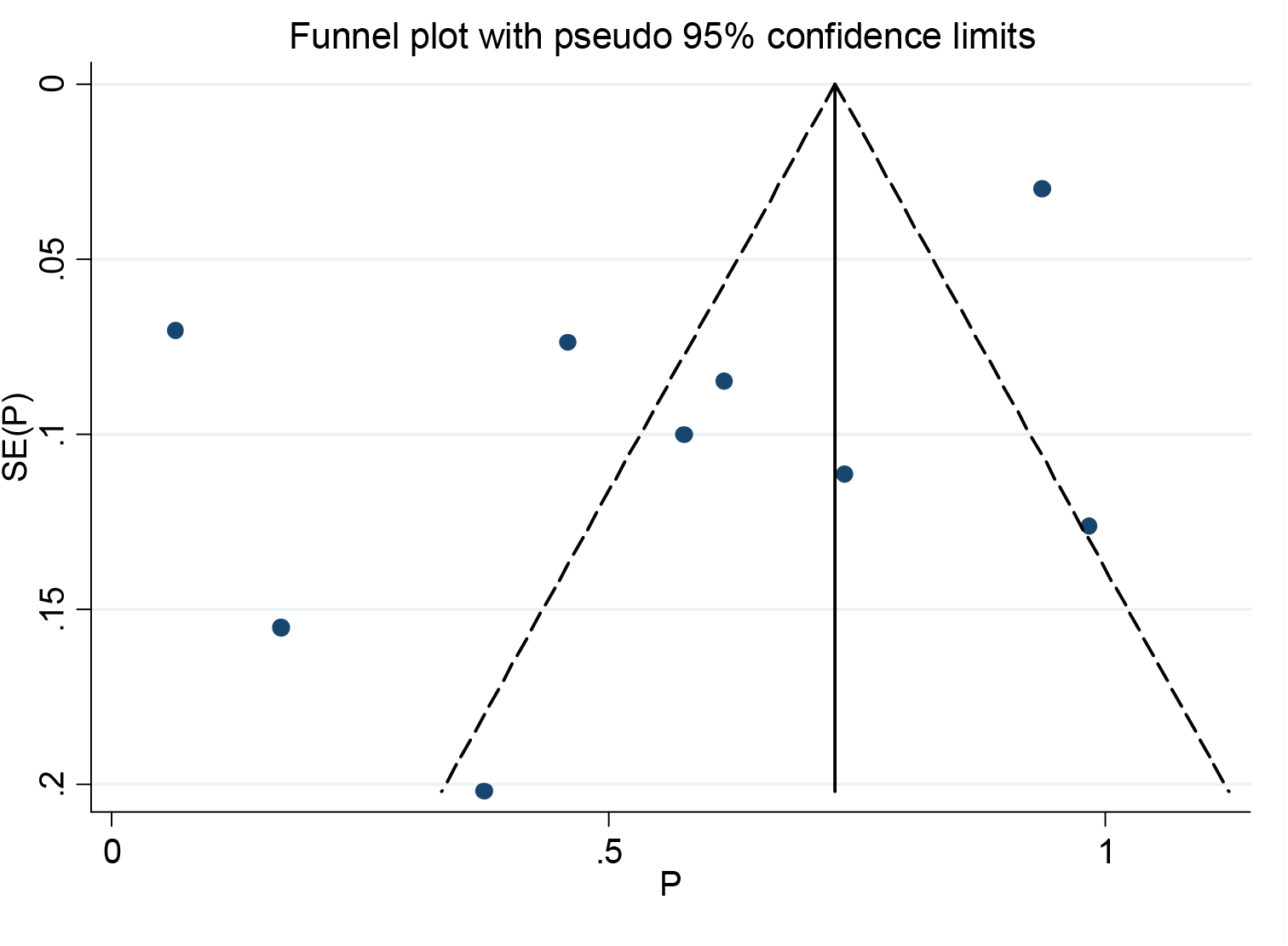
Funnel plot of remain in hospital proportion

## Discussion

Additional to the rapid progression, in late-stage, COVID-19 develop to the ARDS, which is more severe than ARDS occurred routinely (21). The absence of a specific laboratory panel faced the early diagnosis of COVID-19 with a big challenge. Hence there is a critical need to a laboratory interpretation for disease management. In the current survey we evaluated 12 studies, all of them were conducted in China.

Our result showed that the infection is more prevalent in comorbidities situation. Among this hypertension, diabetic, CVD were more common (Table 5). The most observed clinical manifestations were fever, cough, and fatigue (Table 4). Another more common clinical finding was diarrhea, et al. reported a case without COVID-19 clinical characteristics and laboratory abnormality but the virus was detected in stool exam; this show that diarrhea without laboratory abnormality should consider for follow up (22).

COVID-19 entering the alveolar epithelial cells thought angiotensin-converting enzyme 2 (ACE2) (22,23). The main mechanism for inflammation and organ damage is storm cytokines, specially in pulmonary vascular endothelial cells. Hung et al. findings showed that the increasing inflammatory cytokines (IL1B, IL-6, IL-12, IL-10, INF-γ, AND MCP-1) and are higher in ICU admitted, which is a more aggravated disease (20). Increasing inflammation cytokines induction increasing neutrophil count, infiltration them into lung cells, and promoting ARDS developing (12), so elevating neutrophil count should be consider for protecting lung injury. The elevating CRP and ESR are the result of these storm inflammatory cytokines. Additionally, Hung et al. showed that in COVID-19 infected patients, the inhibitory cytokines, like IL-4 and IL-10 increasing, are the leading causes of erythropoiesis inhabitation and lymphopenia in infected patients (20). Our results are in line with this and indicated increasing CRP and ESR, and lymphocytopenia have to consider for COVID-19 infection (Table 3). It is worth to note the Zahng et al. found there is a correlation between eosinopenia and lymphocytopenia, this can be a useful diagnostic marker for COVID-19 diagnosis (9).

Abnormality in liver laboratory tests (e.g., AST, ALT, LDH, bilirubin) was observed in % studies (Table). COVID-19 has receptors on the surface of bile, and these abnormalities can as a result of bile injury (15,23,24). This can be an explanation for normal liver laboratory findings in the early stage of infection. Hypoxia is a serious event in COVID-19, which is one of the main causes of sudden death in patients, hence the increasing CK may be as a result of hypoxia and must be causally interpretation (9).

In a study conducted by Tang et al., it was demonstrated with disease progression, coagulation parameters were increasing; this is in regard with Zhang and Wang et al. findings; that increasing D-dimer and FDP more seen in ICU and severe patients (8–10). Inflammatory cytokines cause activation of monocyte, endothelial cells, expression tissue factor, secretion von Willebrand and finally developing disseminated intravascular coagulation (DIC) (8). It was shown in SARS infection, dysregulation of urokinase pathway, causes activating fibrinolysis and increasing FDPs, and are associated with poor prognosis (8,25).

## Conclusion

In conclusion, the result of the current study showed that in the early stage of COVID-19 infection, maybe there are not significant laboratory findings, but with disease progression, the one or more than signs include increasing AST, ALT, LDH, CK, CRP, ESR, WBC, neutrophil, and decreasing Hemoglobin, lymphocyte count, eosinophil count can be seen. Elevating D-dimer and FDP are associated with ARDS development and can be used as prognostic factors.

## Data Availability

All data presented in manuscript.

## Acknowledgments

We wish to thank all our colleagues in Allied Health Sciences School, Ahvaz Jundishapur University of Medical Sciences.

## Authors’ Contributions

F.R conceived the manuscript, revised it. M.A wrote the manuscript and A.S.M analyzed data and prepared tables and figures.

